# Public Mobility Data Enables COVID-19 Forecasting and Management at Local and Global Scales

**DOI:** 10.1101/2020.10.29.20222547

**Authors:** Cornelia Ilin, Sébastien Annan-Phan, Xiao Hui Tai, Shikhar Mehra, Solomon Hsiang, Joshua E. Blumenstock

## Abstract

Policymakers everywhere are working to determine the set of restrictions that will effectively contain the spread of COVID-19 without excessively stifling economic activity. We show that publicly available data on human mobility — collected by Google, Facebook, and other providers — can be used to evaluate the effectiveness of non-pharmaceutical interventions and forecast the spread of COVID-19. This approach relies on simple and transparent statistical models, and involves minimal assumptions about disease dynamics. We demonstrate the effectiveness of this approach using local and regional data from China, France, Italy, South Korea, and the United States, as well as national data from 80 countries around the world.

**Summary:** *Background:* Policymakers everywhere are working to determine the set of restrictions that will effectively contain the spread of COVID-19 without excessively stifling economic activity. In some contexts, decision-makers have access to sophisticated epidemiological models and detailed case data. However, a large number of decisions, particularly in low-income and vulnerable communities, are being made with limited or no modeling support. We examine how public human mobility data can be combined with simple statistical models to provide near real-time feedback on non-pharmaceutical policy interventions. Our objective is to provide a simple framework that can be easily implemented and adapted by local decision-makers.

*Methods:* We develop simple statistical models to measure the effectiveness of non-pharmaceutical interventions (NPIs) and forecast the spread of COVID-19 at local, state, and national levels. The method integrates concepts from econometrics and machine learning, and relies only upon publicly available data on human mobility. The approach does not require explicit epidemiological modeling, and involves minimal assumptions about disease dynamics. We evaluate this approach using local and regional data from China, France, Italy, South Korea, and the United States, as well as national data from 80 countries around the world.

*Findings:* We find that NPIs are associated with significant reductions in human mobility, and that changes in mobility can be used to forecast COVID-19 infections. The first set of results show the impact of NPIs on human mobility at all geographic scales. While different policies have different effects on different populations, we observed total reductions in mobility between 40 and 84 percent. The second set of results indicate that — even in the absence of other epidemiological information — mobility data substantially improves 10-day case rates forecasts at the county (20.75% error, US), state (21.82 % error, US), and global (15.24% error) level. Finally, for example, country-level results suggest that a shelter-in-place policy targeting a 10% increase in the amount of time spent at home would decrease the propagation of new cases by 32% by the end of a 10 day period.

*Interpretation:* In rapidly evolving disease outbreaks, decision-makers do not always have immediate access to sophisticated epidemiological models. In such cases, valuable insight can still be derived from simple statistic models and readily-available public data. These models can be quickly fit with a population’s own data and updated over time, thereby capturing social and epidemiological dynamics that are unique to a specific locality or time period. Our results suggest that this approach can effectively support decision-making from local (e.g., city) to national scales.

## Introduction

Societies and decision-makers around the globe are deploying unprecedented non-pharmaceutical interventions (NPIs) to manage the COVID-19 pandemic. These NPIs have been shown to slow the spread of COVID-19 (Chinazzi et al., 2020; Ferguson et al., 2020; Hsiang et al., 2020; Tian et al., 2020), but they also create enormous economic and social costs (for example, Atkeson, 2020; Coibion, Gorodnichenko and Weber, 2020; Gössling, Scott and Hall, 2020; Rossi et al., 2020; Thunström et al., 2020). Thus, different populations have adopted wildly different containment strategies (Cheng et al., 2020), and local decision-makers face difficult decisions about when to impose or lift specific interventions in their community. In some contexts, these decision-makers have access to state-of-the-art models, which simulate potential scenarios based on detailed epidemiological models and rich sources of data (for example, Friedman et al., 2020; Ray et al., 2020).

In contrast, many local and regional decision-makers do not have access to state-of-the-art epidemiological models, but must nonetheless manage the COVID-19 crisis with the resources available to them. With global public health capacity stretched thin by the pandemic, thousands of cities, counties, and provinces — as well as many countries — lack the data and expertise required to develop, calibrate, and deploy the sophisticated epidemiological models that have guided decision-making in regions with greater modeling capacity (Gnanvi, Kotanmi et al., 2020; Liverani, Hawkins and Parkhurst, 2013; Loembé et al., 2020). In addition, early evidence suggests a need to adapt models to a local context, particularly for developing countries, where disease, population and other characteristics are different from developed countries, where models are primarily being developed (Evans et al., 2020; Mueller et al., 2020; Twahirwa Rwema et al., 2020).

Here, we aim to address this “modeling-capacity gap” by developing, demonstrating, and testing a simple approach to forecasting the impact of NPIs on infections. This approach is built on two main insights. First, we show that passively collected data on human mobility, which has previously been used to measure NPI compliance (Engle, Stromme and Zhou, 2020; Klein et al., 2020; Kraemer et al., 2020; Martín-Calvo et al., 2020; Morita, Kato and Hayashi, 2020; Pepe et al., 2020; Wellenius et al., 2020), can also effectively forecast the COVID-19 infection response to NPIs up to 10 days in the future. Second, we show that basic concepts from econometrics and machine learning can be used to construct these 10-day forecasts, effectively emulating the behavior of more sophisticated epidemiological models.

This approach is not a substitute for more refined epidemiological models. Rather, it represents a practical and low-cost alternative that may be easily adopted in many contexts when the former is unavailable. It is designed to enable any individual with access to standard statistical software to produce forecasts of NPI impacts with a level of fidelity that is practical for decision-making in an ongoing crisis.

## Data

Our study links information on non-pharmaceutical interventions (NPIs, shown in Figure 1a) to patterns of human mobility (Figure 1b) and COVID-19 cases (Figure 1c-d). All data were obtained from publicly available sources. We provide a brief summary of these data here; full details are provided in Appendix A.

**Figure 1:**
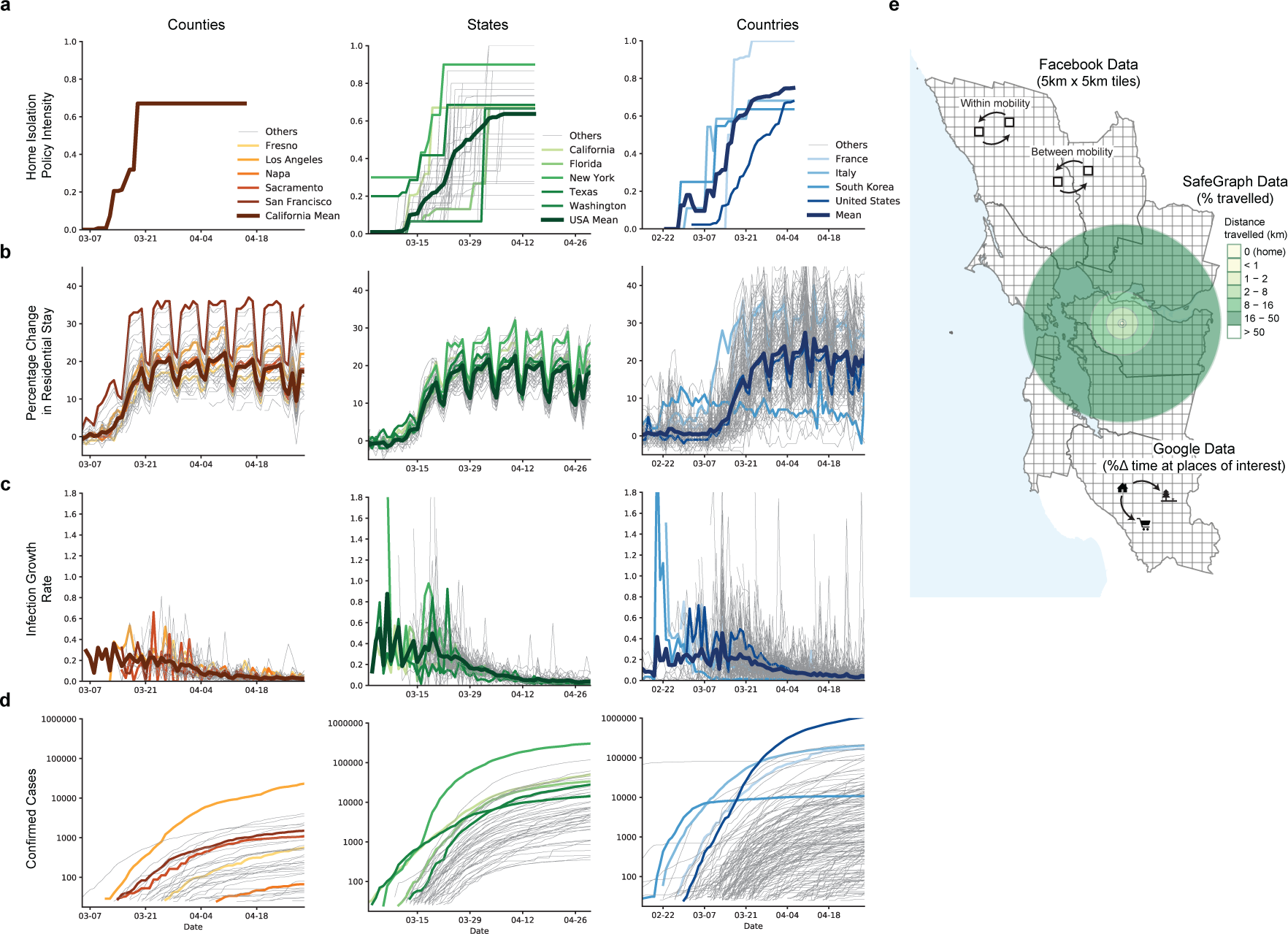
Data on mobility measures, COVID-19 infections and home isolation policy adoption. (**a**) Home isolation policy adoption, (**b**) Change in time spent at home, (**c**) Infection growth rate, and (**d**) Total confirmed cases are displayed at the county, state and country level. (**e**) Illustrative example of different mobility measures in California. We utilize data on trips both within and between counties (Facebook and Baidu) as well as the purpose of the trip (Google) and the average distance traveled (SafeGraph).

### Non-Pharmaceutical Interventions

We obtain NPI data from two sources. At the sub-national level, we use the NPI dataset compiled by Hsiang et al. (2020).^1^ For each sub-national region in five countries, we observe the fraction of the population treated with NPIs in each location on each day. We aggregate 13 different policy actions into four general categories: Shelter in Place, Social Distance, School Closure, and Travel Ban. At the national level, we compiled data on national lockdown policies from the Organisation for Economic Co-operation and Development (OECD) - Country Policy Tracker,^2^ and crowed-sourced information on Wikipedia and COVID-19 Kaggle competitions.^3^

### Human Mobility

We source publicly-available data on human mobility from Google, Facebook, Baidu and SafeGraph. These private companies provide free aggregated and anonymized information on the movement of users of their online platform (Fig 1e). Data from Google indicates the percentage change in the amount of time people spend in different types of locations (e.g., residential, retail, and workplace).^4^ These changes are relative to a baseline defined as the median value, for the corresponding day of the week, during Jan 3–Feb 6, 2020. Facebook provides estimates of the number of trips within and between square tiles (of resolution up to 360m^2^) in a region.^5^ We aggregate these data to show trips between and within sub-national units. Baidu provides similar data, indicating movement between and within major Chinese cities.^6^ Lastly, SafeGraph dataset gives us information on average distance travelled from home by millions of devices across the US.^7^

### COVID-19 Cases

For each subnational and national unit, we obtain the cumulative confirmed cases of COVID-19 from the data repository compiled by the Johns Hopkins Center for Systems Science and Engineering (JHU CSSE COVID-19 Data).^8^

### Linking Data Sets

The availability of epidemiological, policy, and mobility data varies across subnational units and countries included in the analysis. We distinguish between three different levels of aggregation for administrative regions - denoted “ADM2” (the smallest unit), “ADM1”, “ADM0.” Our global analysis is conducted using ADM0 data. The country-specific analysis is determined by data availability. Results are provided at the prefecture (ADM2) and province level (ADM1) in China; the regional (ADM1) level in France; the province (ADM2) and region (ADM1) level in Italy; the province (ADM1) level in South Korea; and the county (ADM2) and state (ADM1) level in the United States.

We merge the sub-national NPI, mobility, and epidemiological data based on administrative unit and day to form a single longitudinal (panel) data set for each country. We merge the daily country-level observations to construct a longitudinal data sets for the portion of the world we observe.

## Methods

### Models

We decompose the impact of an NPI on infections 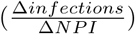 into two components that can be modeled separately: the change in behavior associated with the NPI, and the resulting change in infections associated with that change in behavior:

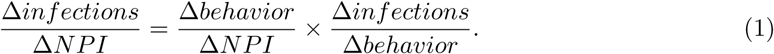

We construct models to describe each of these two factors. The “behavior model” describes how mobility behavior changes in association with the deployment of NPIs 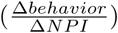. The “infection model” describes how infections change in association with changes in mobility behavior 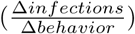. Both models are “reduced-form” models, commonly used in econometrics, that characterize the behavior of these variables without explicitly modeling the underlying mechanisms that link them (cf., Hsiang et al., 2020). Instead, these models emulate the output one would expect from more sophisticated and mechanistically explicit epidemiological models — without requiring the underlying processes to be specified. While this reduced-form approach does not provide the same epidemiological insight that more detailed models do, they demand less data and fewer assumptions. For example, they can be fit to local data by analysts with basic statistical training, not necessarily in epidemiology, and they do not require knowledge of fundamental epidemiological parameters — some of which may differ in each context and can be difficult to determine. The performance of these simple, low-cost models can then be evaluated via cross-validation, i.e., by systematically evaluating out-of-sample forecast quality.

### Behavior Model

For each country, we separately estimate how daily sub-national mobility behavior changes in association with the deployments of NPIs using a country-specific model. In the global model, we pool data across countries and estimate how mobility in each country changes in association with national exposure to NPIs. Each category of mobility on each day is assumed to be simultaneously influenced by the collection of NPIs that are active in that location on that day. A panel multiple linear regression model is used to estimate the relative association of each category of mobility with each NPI. Our approach accounts for constant differences in baseline mobility between and within each sub-national unit – such as differences due to regional commuting patterns, culture, or geography, and differences in mobility across days of the week. These effects are not modeled explicitly but instead are accounted for non-parametrically. Appendix B.1 contains details of the modeling approach.

### Infection Model

As with the behavior model, we model the daily growth rate of infections at the local, national, and global scale. In each location, we model the daily growth rate of infections as a function of recent human mobility and historical infections. The approach does not require epidemiological parameters, such as the incubation period or *R*_0_, nor information on NPIs.

In practice, we estimate a distributed-lag model where the predictor variables are mobility rates in that location for the prior 21 days, and the dependent variable is the daily infection growth rate, constructed as the first-difference of log confirmed infections. This approach captures the intuition that human mobility is a key factor in determining rates of infection, but does not require parametric assumptions about the nature of that dependency. The model also accounts for constant differences in baseline infection growth rates within each locality — such as those due to differences in local behavior unrelated to mobility, differences across days of the week, and changes in how confirmed infections are defined or tested for. This approach is also robust to incomplete rates of COVID-19 testing, uneven patterns of testing across space, and gradual changes in testing over time (Hsiang et al., 2020).

We fit the model using historical data from each location, and follow stringent practices of cross-validation to ensure that the models are not ‘overfit’ to historical trends. The accuracy of the forecast is then evaluated against actual infections observed during the forecast period, but which were not used to fit the model. Models are fit at the finest administrative level where data are available and forecasts are aggregated to larger regions to evaluate the ability of the model to predict infections at different spatial scales. Appendix B.2 contains details of the modeling approach.

In principle, such future forecasts can be used by decision-makers who are able to influence local mobility through policy and/or NPIs, perhaps informed either by a behavioral model or observation. Here, we test the quality of the infection model to generate forecasts by simulating and evaluating what a forecaster would have predicted had they generated a forecast at a historical date. In the forecasts presented here, we assume that mobility remains at the level observed during the forecast period – although in practice we expect that decision-makers would simulate different forecasts under different mobility assumptions to inform NPI deployment and policy-making.

## Results

We first present results from our behavior model, characterizing the mobility response of different populations to different NPIs. We then evaluate the infection model’s ability to forecast COVID-19 infections based on these same mobility measures. We conclude by discussing how these models could be used to guide policy decisions at local and regional scales.

### Mobility response to NPIs

We estimate the reduction in human mobility associated with the deployment of NPIs by linking comprehensive data on policy interventions to mobility data from several different countries at multiple geographic scales. We find that the combined impact of all NPIs reduced mobility between administrative units (Facebook/Baidu) by 73% on average across the countries with sub-national policy data (Fig 2a). The combined effects were of similar magnitude in China (−78%, se = 8%), France (−88%, se = 27%), Italy (−85%, se = 12%), and the US (−69%, se = 6%); no significant change was observed in South Korea, where mobility was not a direct target of NPIs (for example, You, 2020). Excluding South Korea, we estimate that all policies combined were associated with a decrease in mobility by 81%. The general consistency of these magnitudes across countries holds for alternative measures of mobility: using Google data we find that all NPIs combined result in an increase in time spent at home by 28% (se = 2.9), 24% (se = 1.3), and 26% (se = 1.3) in France, Italy, and the US, respectively. This was achieved, in part, by reducing time spent at workplaces by an average of 59.8% and time in commercial retail locations by an average of 78.8%.

**Figure 2:**
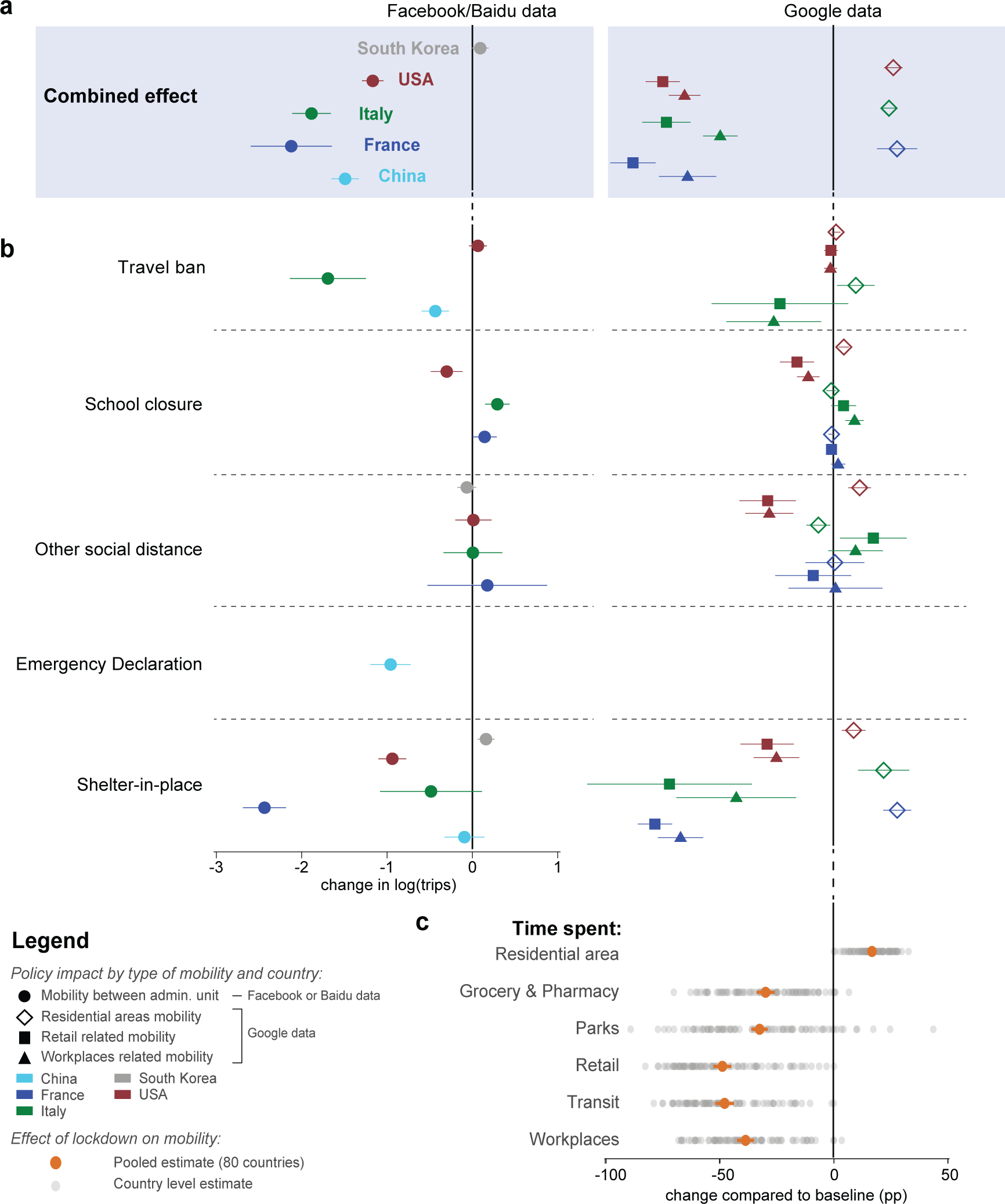
Empirical estimates of the effect of NPIs on mobility measures. Markers are country specific-estimates, whiskers show the 95% confidence interval. **a** Estimated combined effect of all policies on number of trips between counties (left) and time spent in specific places (right). **b** Estimated effects of individual policy or policy groups on mobility measures, jointly estimated for each country. **c** Estimated effect of lockdown on mobility the 80 countries which experienced such policy, jointly estimated for each type of mobility.

We estimate the impact of each individual NPI on total trips (Facebook/Baidu) and quantity of time spent at home and other locations (Google) accounting for the estimated impact of all other NPIs. Travel bans are significantly associated with large mobility reductions in China (−70%, se = 7%) and Italy (−82%, se = 25%), where individuals stayed home for 10% more time, but not in the US (Fig 2b). School closures were associated with moderate negative impacts on mobility in the US (−26%, se = 10%) and increased time at home (4.6%, se = 0.7%) but slight positive impacts in Italy (33%, se = 7%) and France (15%, se = 7%). Other social distancing policies, such as religious closures, had no consistent impact on total trips but were associated with individuals spending more time at home in the US (11.5%, se = 1.6%) and more time in retail locations in Italy (17.6%, se = 4.8%). Similarly, the national emergency declaration was associated with significant mobility reductions in China (−62.6 %, se = 12.7 %). Shelter-in-place orders were associated with large reductions in trips for the US (−60.8%, se = 8%), Italy (−38.4%, se = 35%), and France (−91.2%, se = 13.6%), and large increases in the fraction of time spent in homes (8.9%, 22.1%, 28%, respectively). Shelter in place orders did not appear to have large impacts in South Korea or China. This is consistent with earlier policies (such as the Emergency Declaration) restricting movement in China earlier than the shelter in place orders, while mobility in South Korea was never substantially affected by NPIs.

Globally, we find evidence that lockdown policies were associated with substantial reductions in mobility (Figure 2c). Across 80 countries, the average time spend in non-residential locations decreased by 40% (se = 2%) in response to NPIs. Time spent in retail locations is the most impacted category, declining 49.9% (se = 2%). Some of the variation in response across countries (grey dots) likely reflects different social, cultural, and economic norms; measurement error; and statistical variability. In Appendix C, we disaggregate this effect temporally, and find that the most significant reductions occur during the first eight days after a lockdown (Figure 5c).

In Appendix C, we further exploit the granular resolution of the mobility data to investigate whether localized policies also impacted neighboring regions (Figure 5). In the USA and Italy, the impact of NPIs on mobility was highly localized, with little evidence of spatial spillover effects (Appendix C - Figure 5a). In China, the evidence is more mixed, with some evidence of spillovers between neighboring cities (Appendix C - Fig 5b).

### Forecasting infections based on mobility

We find that mobility data alone are sufficient to meaningfully forecast COVID-19 infections 7-10 days ahead at all geographic scales – from counties and cities (ADM2), to states and provinces (ADM1), to countries (ADM0) and the entire world. Furthermore, identical models that exclude mobility data perform substantially worse, suggesting an important role for mobility data in fore-casting.

Figure 3 illustrates the performance of model forecasts in several geographic regions and at multiple scales. The true infection rate is shown as a solid line; data used to train each model are depicted in blue dots, and the forecast of our model is shown in orange, contrasted against a model with no mobility data in green. Forecasts that account for current and lagged measures of mobility generally track actual cases more closely than forecasts that do not account for mobility.

**Figure 3:**
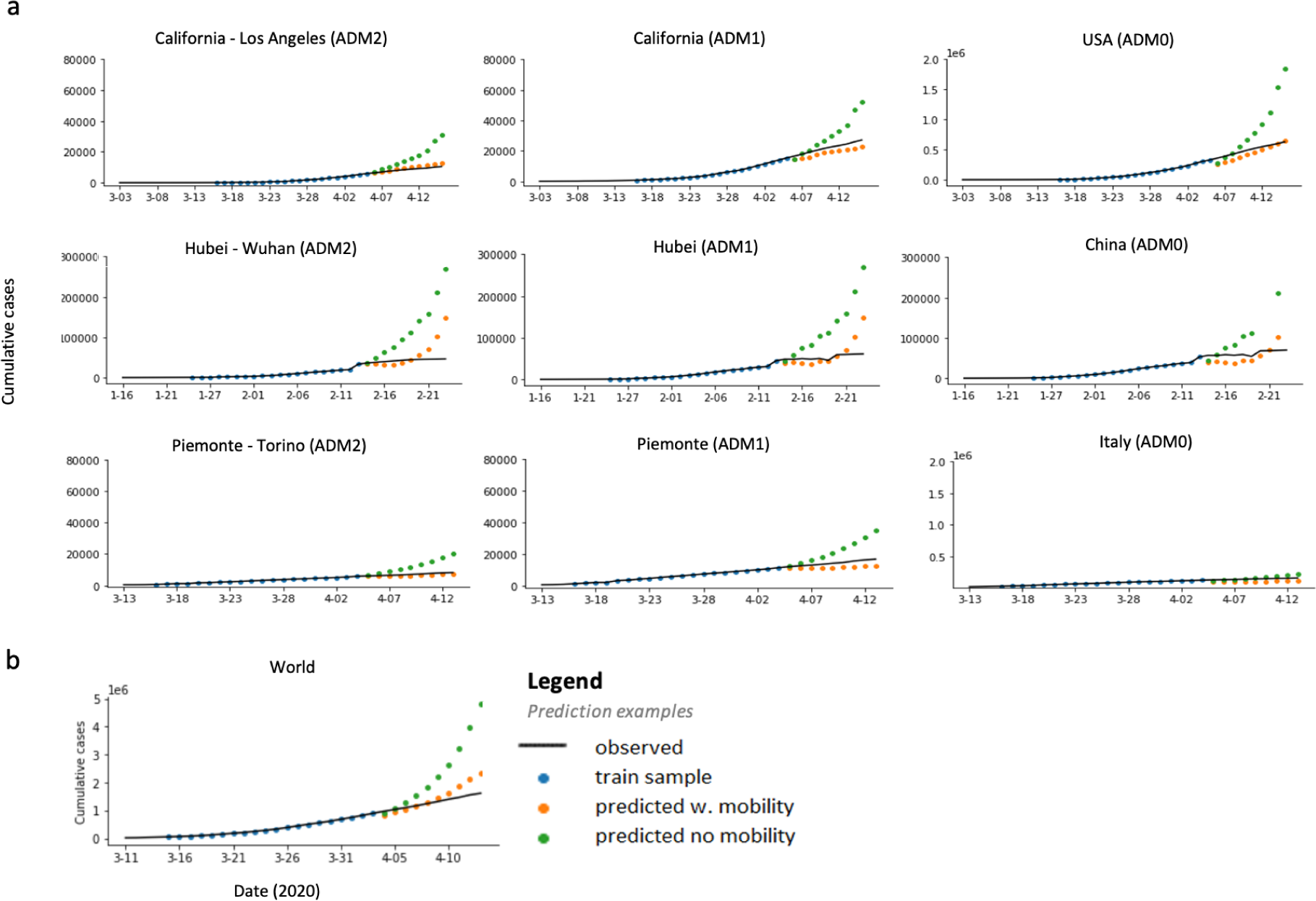
Short term prediction of COVID-19 cases. Solid line is the recorded number of COVID-19 infections, markers show data in our training sample (blue) and our predictions estimated using mobility measures (orange) versus a model without mobility (green). Model with no mobility measures consistently over-predict the number of infections and drift away quickly from the observed data. **a** This pattern is confirmed when aggregating locally estimated predictions (left) at the state (middle) and country (right) level. **b** Similarly, predictions obtained from country level estimates are significantly more accurate when a measure of mobility is included.

For example, a forecast made for the period 4*/*06*/*2020 − 4*/*15*/*2020 for California-Los Angeles on 4/15/2020 without mobility projects 30, 716 cases, while the same forecast accounting for mobility would be 12, 650 cases, much closer to the 10, 496 that was observed. Figure 3b depicts projected cases for the entire world based on this reduced-form approach, estimated using country-level data mobility data from Google.

Figure 4 summarizes model performance across *all* administrative subdivisions of each of the three countries we consider for the forecast analysis (China, Italy, and the United States). We show the distribution of model errors over all ADM2 and ADM1 regions at forecast lengths ranging from 1 to 10 days. Table 1 summarizes each distribution using the median.

**Table 1:** Median percentage error for each model and day of forecast, as plotted in Figure 4. The error is presented for each model and geographical region, and for 1 to 10 day forecasts.

| Model | Country | Level | Median percentage error (MPE) for forecast length (days) |  |  |  |  |  |  |  |  |  |
| --- | --- | --- | --- | --- | --- | --- | --- | --- | --- | --- | --- | --- |
|  |  |  | 1 | 2 | 3 | 4 | 5 | 6 | 7 | 8 | 9 | 10 |
| Mobility | World | ADM0 | 0.90 | 2.19 | 3.46 | 4.83 | 6.35 | 7.80 | 9.54 | 11.00 | 12.60 | 15.24 |
| No Mobility | World | ADM0 | 1.50 | 3.84 | 6.32 | 8.70 | 11.46 | 14.42 | 16.91 | 19.75 | 25.28 | 31.12 |
| Mobility | China | ADM1 | -0.20 | 1.41 | 3.05 | 4.55 | 5.91 | 7.54 | 7.60 | 17.59 | 53.26 | 124.82 |
| No Mobility | China | ADM1 | 1.47 | 5.39 | 8.84 | 14.10 | 18.45 | 23.45 | 26.80 | 45.86 | 64.95 | 112.03 |
| Mobility | China | ADM2 | -0.44 | 0.89 | 2.11 | 3.29 | 4.18 | 5.19 | 6.10 | 6.80 | 50.66 | 131.09 |
| No Mobility | China | ADM2 | 1.37 | 5.17 | 8.59 | 12.48 | 16.83 | 22.33 | 28.14 | 35.70 | 65.09 | 128.80 |
| Mobility | Italy | ADM1 | 0.89 | 2.70 | 5.60 | 5.68 | 8.29 | 6.65 | 9.31 | 6.83 | 14.39 | 34.19 |
| No Mobility | Italy | ADM1 | 4.86 | 13.66 | 23.12 | 33.69 | 45.03 | 61.06 | 80.95 | 107.48 | 133.98 | 172.74 |
| Mobility | Italy | ADM2 | -0.87 | -0.90 | -0.81 | -1.06 | -1.73 | -2.11 | -2.08 | -0.99 | 4.41 | 13.27 |
| No Mobility | Italy | ADM2 | 4.96 | 14.20 | 23.75 | 35.26 | 45.81 | 61.66 | 73.96 | 95.47 | 128.89 | 167.97 |
| Mobility | US | ADM1 | 1.13 | 2.62 | 3.97 | 5.45 | 6.68 | 8.52 | 11.19 | 14.28 | 17.24 | 21.82 |
| No Mobility | US | ADM1 | 3.50 | 9.39 | 15.29 | 21.79 | 29.22 | 37.11 | 45.54 | 57.36 | 71.72 | 88.73 |
| Mobility | US | ADM2 | 0.98 | 2.30 | 3.64 | 5.21 | 7.00 | 8.77 | 11.00 | 14.28 | 16.67 | 20.75 |
| No Mobility | US | ADM2 | 2.80 | 7.72 | 12.62 | 17.79 | 23.79 | 30.34 | 37.37 | 48.43 | 63.31 | 79.47 |

**Figure 4:**
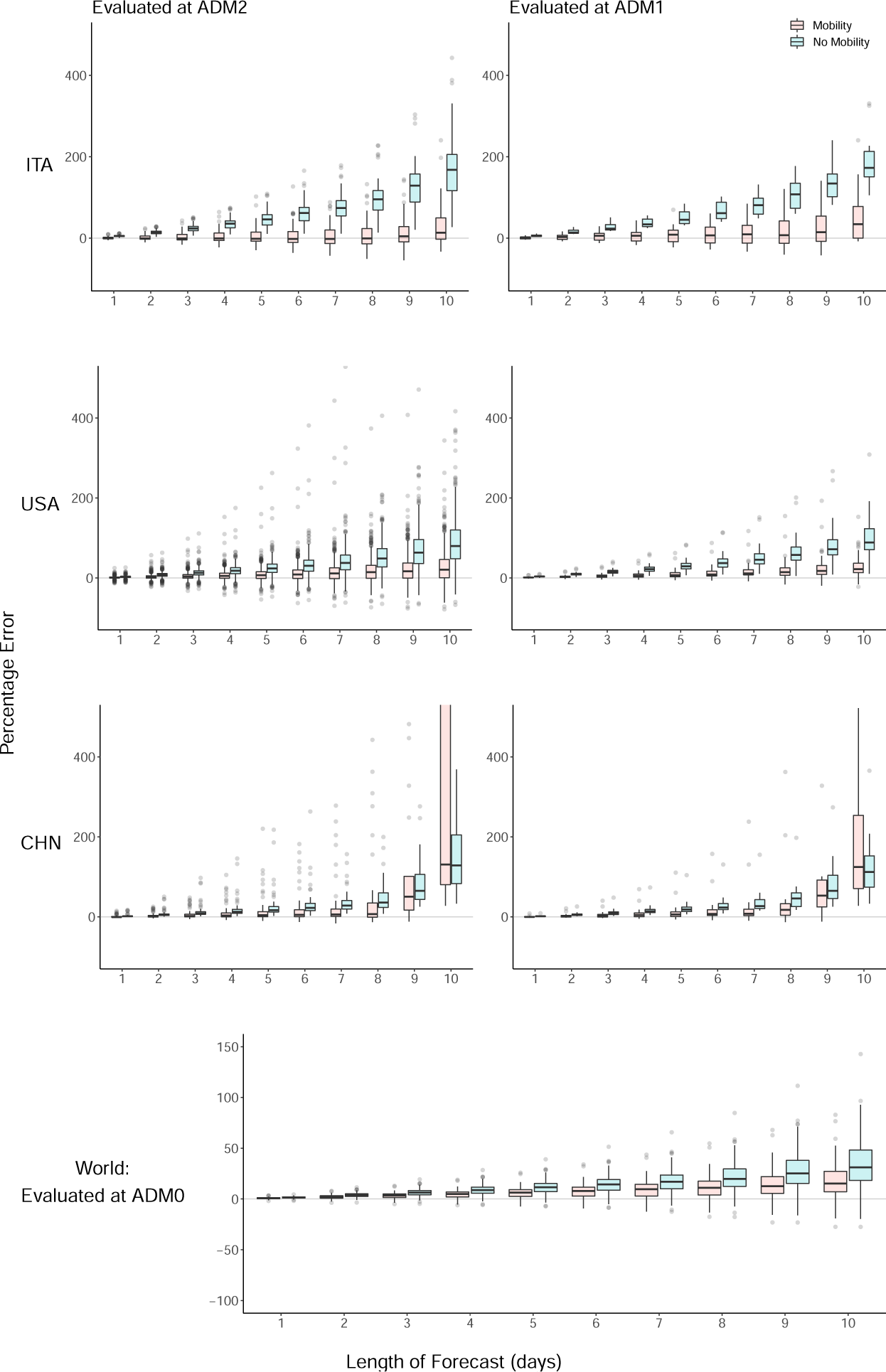
Evaluation of forecast errors for the infection model. For Italy, US and China, forecasts are evaluated at the finest administrative level (ADM2), as well as aggregated to larger regions (ADM1). For each ADM2 region and forecast length, the mean is taken over all available forecast dates, and the error is evaluated using that mean. Boxplots display the distribution of these percentage errors for each ADM2 region. These are then aggregated to ADM1 level (right panel), for both models including and excluding mobility variables. Similarly, for data fitted at a global level (bottom-most plot), for each country and forecast length, the mean is taken over all forecast dates.

**Figure 5:**
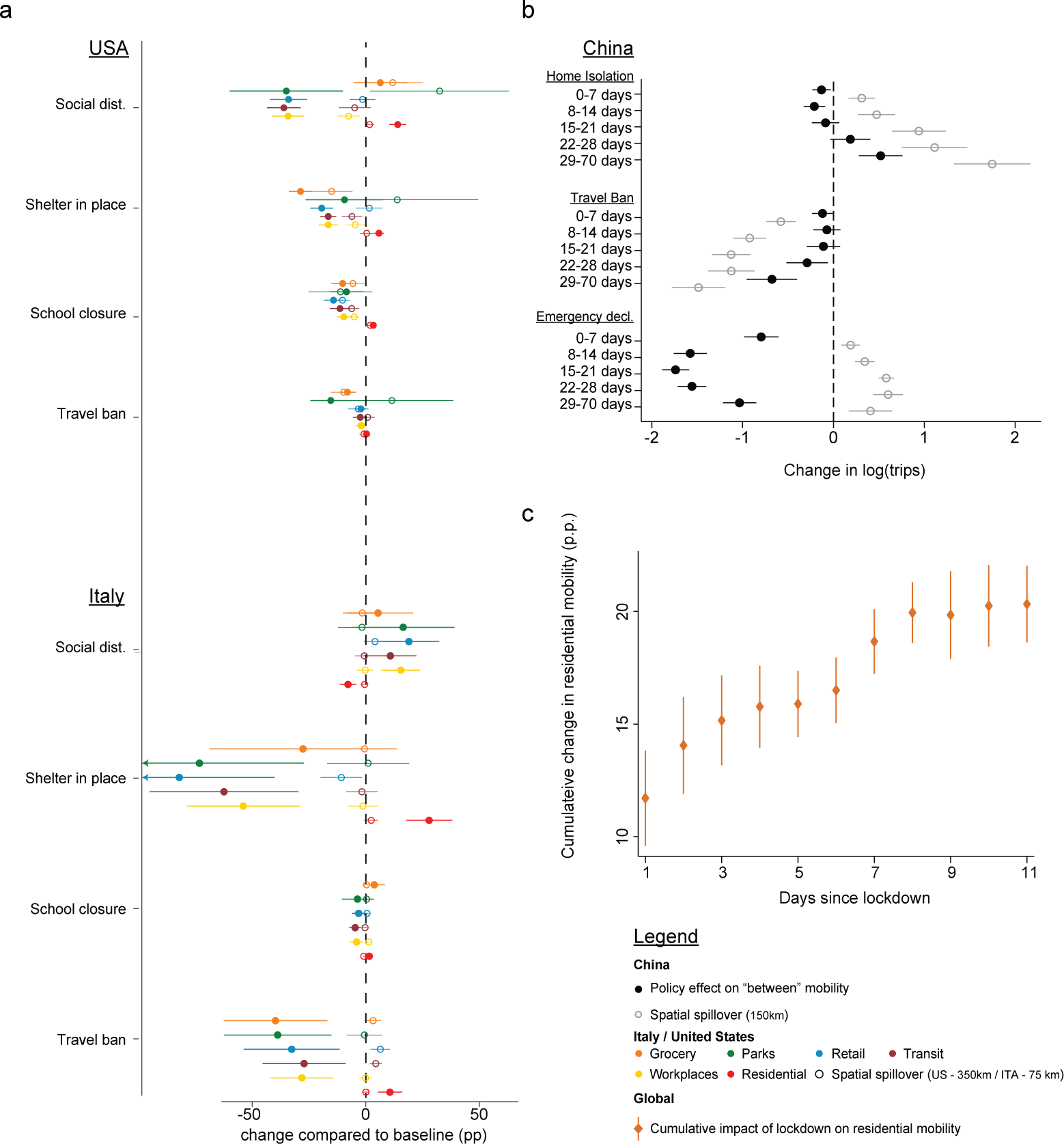
Spatial and temporal spillover of policies. **a-b.** Solid markers indicate the direct impact of large policies on mobility. Hollow markers show the estimated effect of a policy on neighboring regions. Policies are jointly estimated at the local level for each country. In China (**b**), we also separately estimate the effect of each policy for each time period after the policy’s implementation. **c** The impact of lockdown on the time spent at home is estimated using a countrylevel regression with 80 countries. We report the cumulative effect over time.

In all geographies and at all scales, models with mobility data perform better than models without. In general, sub-national forecasts in China benefit least from mobility data, but forecasts in Italy and the US are substantially improved by including a single measure of mobility for the 21 days prior to the date of the forecast. At the local (ADM2) level in Italy, the MPE is -1.73% and 13.27% for five and ten days in the future when mobility is accounted for, compared to 45.81% and 167.97% when it is omitted. In the US, MPE is 7.00% (5-day) and 20.75% (10-day) accounting for mobility, and 23.79% and 79.47% omitting mobility. In China, MPE is 4.18% (5-day) and 131.09% (10-day) accounting for mobility, and 16.83% and 128.80% omitting mobility. At the regional (ADM1) level, MPE rates are similar but extreme errors are reduced, largely because positive and negative errors cancel out. Country-level forecasts, which use country-level mobility data from Google, benefit relatively less than sub-national model from including mobility information, in part because baseline forecast errors are smaller. For countries in our sample, MPE is 6.35% (5-day) and 15.24% (10-day) accounting for mobility, and 11.46% and 31.12% omitting mobility.

### Model application in decentralized management of infections

Our results suggest that a simple reduced-form approach to estimating model (1) may provide useful information and feedback to decision-makers who might otherwise lack the resources to access more sophisticated scenario analysis. We imagine the approach can be utilized in two ways. First, a decision-maker considering an NPI (either deploying, continuing, or lifting) could develop an estimate for how that NPI might affect behavior, based on our analysis of different policies above (Fig 2). Using these estimated changes in mobility, they could then forecast changes in infections using the infection model described above — but fit to local data.

Table 2 provides an example calculation for how a novel policy that increased residential time (observed in Google data) would alter future infections, using estimates from the global-level model. For example, a policy that increases residential time by 5% in a country is predicted to reduce cumulative infections ten days later, to 82.5% (CI: (78.2, 87.0)) of what they would otherwise have been. Similar tabulations can be generated by fitting infection models using recent and local data, which would flexibly capture local social, economic, and epidemiological conditions.

**Table 2:** Example: Estimating the effect of a mobility-reducing policy on infections for the global model (unit of observation is a country day). The values in the table are the ratio of the cumulative number of cases after up to 10 days, if residential time over baseline was increased in a country *at day 0* by Δ = 1%, 5%, 10% or 20% from their original values. These values are estimated using coefficients of the mobility variables derived from the pooled global model (details in Appendix B.2). Note that each column compares to the value on its first row (indicated by the value 1). An example interpretation is: if a country increases residential time by 5%, cumulative infections ten days later is predicted to be 82.5% of what they would have been with no change in mobility.

| $\Delta$ | day = 1 | 2 | 3 | 4 | 5 | 6 | 7 | 8 | 9 | 10 |
| --- | --- | --- | --- | --- | --- | --- | --- | --- | --- | --- |
| 0 | 1 | 1 | 1 | 1 | 1 | 1 | 1 | 1 | 1 | 1 |
| .01 | 1 | 0.998 | 0.996 | 0.993 | 0.989 | 0.985 | 0.98 | 0.975 | 0.969 | 0.962 |
| .05 | 0.998 | 0.992 | 0.981 | 0.966 | 0.948 | 0.928 | 0.906 | 0.881 | 0.854 | 0.825 |
| .10 | 0.996 | 0.983 | 0.962 | 0.934 | 0.899 | 0.862 | 0.821 | 0.776 | 0.729 | 0.68 |
| .20 | 0.992 | 0.967 | 0.926 | 0.873 | 0.808 | 0.743 | 0.674 | 0.603 | 0.532 | 0.463 |

A second way that a decision-maker could use our approach would be to actually deploy a policy without *ex ante* knowledge of the effect it will have on mobility, instead simply observing mobility responses that occur after NPI deployment using these publicly available data sources. Based on these observed responses, they could forecast infections using our behavior model.

## Discussion

The COVID-19 pandemic has led to an unprecedented degree of cooperation and transparency within the scientific community, with important new insights rapidly disseminated freely around the globe. However, the capacity of different populations to leverage new scientific insights is not uniform. In many resource-constrained contexts, critical decisions are not supported by robust epidemiological modeling of scenarios. Here we have demonstrated that freely available mobility data can be used in simple models to generate practically useful forecasts. The goal is for these models to be accessible to a single individual with basic training in regression analysis using standard statistical software. The reduced-form model we develop generally performs well when fit to local data, except in China where it cannot account for some key factors that contributed to reductions in transmission.

A key insight from our work is that passively observed measures of aggregate mobility are useful predictors of growth in COVID-19 cases. However, this does not imply that population mobility itself is the only fundamental cause of transmission. The measures of mobility we observe capture a degree of “mixing” that is occurring within a population, as populations move about their local geographic context. This movement is likely correlated with other behaviors and factors that contribute to the spread of the virus, such as low rates of mask-wearing and/or physical distancing. Our approach does not explicitly capture these other factors — and thus should not be used to draw causal inferences — but is possible that our infection model performs well in part because the easy-to-observe mobility measures capture these other factors by proxy.

The simple model we present here is designed to provide useful information in contexts when more sophisticated process-based models are unavailable, but it should not necessarily displace those models where they are available. In cases where complete process-based epidemiological models have been developed for a population and can be deployed for decision-making, the model we develop here could be considered complementary to those models. Future work might determine how information from combinations of qualitatively distinct models can be used to optimally guide decision-making.

We also note that the reduced-form model is designed to forecast infections in a certain population at a restricted point in time. It achieves this by capturing dynamics that are governed by many underlying processes that are unobserved by the modeler. However, because these underlying mechanisms are only captured implicitly, the model is not well-suited to environments where these underlying dynamics change dramatically. In such circumstances, process-based models will likely perform better. Nonetheless, the reduced-form approach presented here can also be applied in these circumstances, but it may be necessary to refit the model based on data that is representative of current conditions. Similarly, when our reduced-form model is applied to a new population, it should be fit to local data to capture dynamics representative of the new population.

The approach we present here depends critically on the availability of aggregate mobility data, which is currently provided to the public by private firms that passively collect this information. We hypothesize that the approach we develop here might skillfully forecast the spread of other diseases besides COVID-19. If true, this suggests our approach could provide useful information to decisionmakers for managing other public health challenges, such as influenza or other outbreaks, potentially indicating a public health benefit from firms continuing to made mobility data available—even after the COVID-19 pandemic has subsided.

## Data Availability

The data are publicly available and described in the manuscript.

## Acknowledgements

We thank Jeanette Tseng for her role in designing Fig. 1. S.A.P. is supported by a gift from the Tuaropaki Trust. This material is based upon work supported by the National Science Foundation under Grant IIS-1942702. Any opinions, findings, and conclusions or recommendations expressed in this material are those of the authors and do not necessarily reflect the views of the National Science Foundation. Funding was also provided by Award 2020-0000000149 from CITRIS and the Banatao Institute at the University of California. None of the authors has been paid to write this article by a pharmaceutical company or other agency. All authors had full access to the full data in the study and accept responsibility to submit for publication.

## Author contributions

J.B. and S.H. conceived and led the study. C.I., J.B., S.A.P., S.H., X.H.T., designed analysis, and interpreted results. C.I., S.A.P., S.M., and X.H.T. collected, verified, cleaned and merged data. C.I. created Figs. 3, 6 and Table 3. S.A.P. created Figs. 2, 5. S.M. created Fig. 1 and Table 6. X.H.T. created Fig.4 and Tables 1,2. X.H.T. managed literature review. All authors wrote the paper. C.I., S.A.P., and X.H.T. contributed equally and are listed in a randomly assigned order.

**Table 3:** Mobility Data Sources. - Details of mobility data sources for each country. The table provides relevant dates and level of analysis used for the behavior model and infection model, respectively.

| Region | Mobility Data | Level of analysis | Type of mobility | Start Date | End Date |
| --- | --- | --- | --- | --- | --- |
| **Behavior model** |  |  |  |  |  |
| China | Baidu | ADM2 | between | 1/10/2020 | 3/6/2020 |
| France | Facebook | ADM1 | between | 3/4/2020 | 4/8/2020 |
| Italy | Facebook | ADM2 | between | 2/25/2020 | 4/8/2020 |
| South Korea | Facebook | ADM2 | between | 2/23/2020 | 4/6/2020 |
| United States | Google | ADM2 | residential,<br>retail,<br>workplaces | 3/3/2020 | 4/12/2020 |
| World | Google | ADM0 | residential,<br>retail,<br>workplaces | 2/26/2020 | 3/28/2020 |
| **Infection model** |  |  |  |  |  |
| China | Baidu | ADM1, ADM2 | between | 2/12/2020 | 3/3/2020 |
| France | Facebook | - | - | - | - |
| Italy | Facebook | ADM1, ADM2 | between | 3/31/2020 | 4/13/2020 |
| South Korea | Facebook | - | - | - | - |
| United States | Google | ADM1, ADM2 | - | 3/17/2020 | 4/30/2020 |
| World | Google | ADM0 | - | mobility today ≥ 5% | 5/29/2020<br>or until growth rates<br>of new cases flatten |

**Figure 6:**
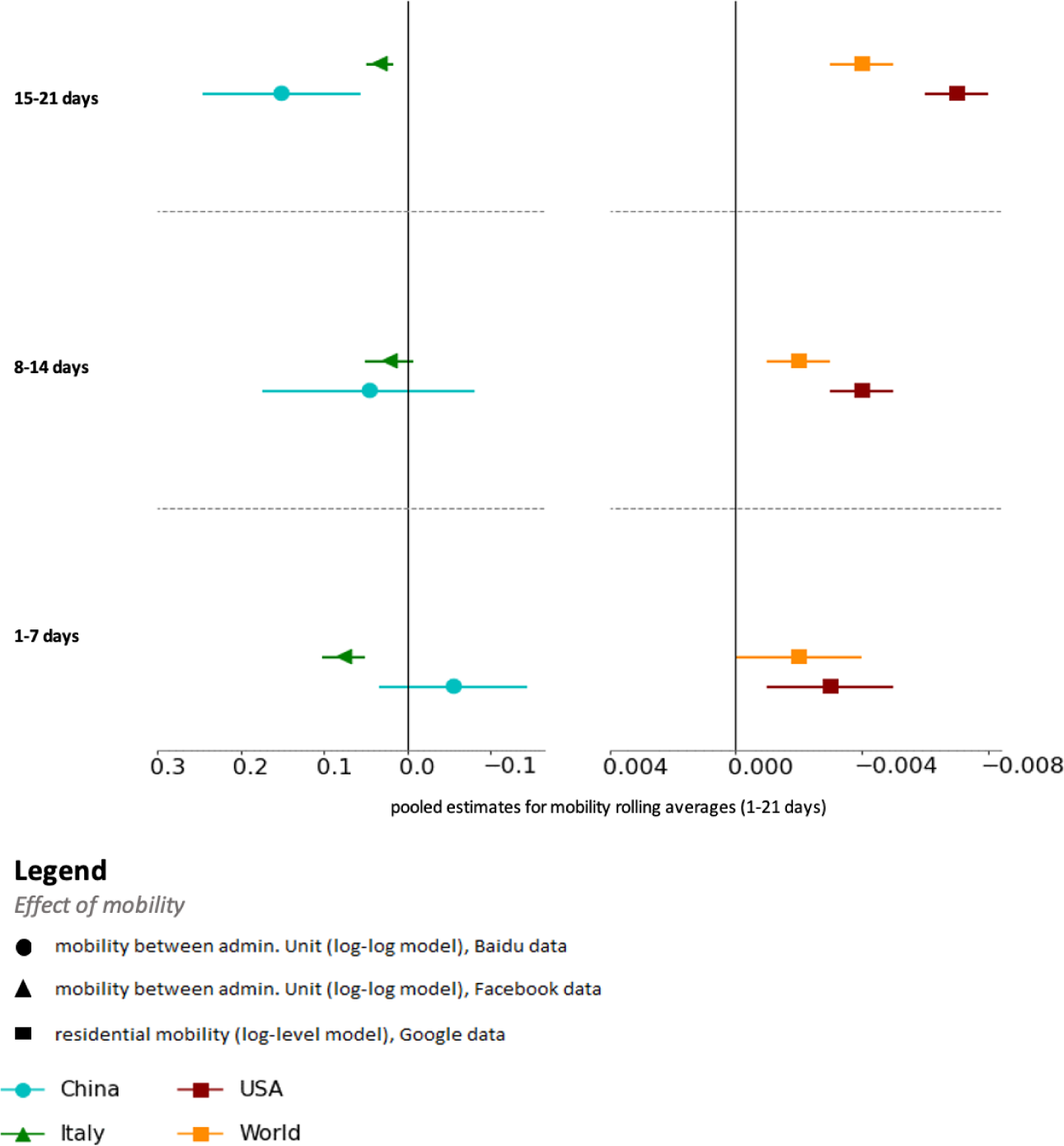
Impact of mobility on the growth rate of COVID-19 cases. Estimated impact of mobility on COVID-19 infection growth rate over time. Effects are estimated for each of the preceding three weeks (lags of 1 to 21 days), where the measure of mobility is either the number of trips between administrative units (left) or the amount of time spent at home (right). The impact of mobility is gradually increasing over time and is highest after 2 weeks.

## Appendices

### A Data Acquisition and Processing

Data used in this study can be divided into three categories - Epidemiological, Policy and Mobility. The sources of these data sets include various research institutions, government public health websites, regional newspaper articles and digital social media platforms.

#### A.1 Epidemiological Data

We collected epidemiological data from the 2019 Novel Coronavirus COVID-19 (2019-nCoV) Data Repository compiled by the Johns Hopkins Center for Systems Science and Engineering (JHU CSSE).^9^ The primary variable of interest for our study is *cum confirmed cases*, i.e., the total number of confirmed positive cases in an administrative area since the first confirmed case. We accessed it along with other relevant metadata, including:

*date*: The date of observation

*adm0 name*: The ISO3 region (Administrative Level 0) code of the observation

*adm1 name*: The name of the “Administrative Level 1” region of the observation

*adm2 name*: The name of the “Administrative Level 2” region of the observation

#### A.2 Policy data

The policy data was constructed and made available for academic research by Hsiang et al. (2020).^10^ For each country, the relevant country-specific policies were identified and mapped to four harmonized policy categories - Travel Ban, School Closure, Shelter in place, and Social Distance. These category variables were created by taking an average of policy variables related to that category.

##### i. Travel Ban

- *travel ban local* : Represents a policy that restricts people from entering or exiting the administrative area (e.g., county or province) treated by the policy.

##### ii. School Closure

- *school closure*: Represents a policy that closes school and other educational services in that area.

##### iii. Shelter In Place

- *home isolation*: Represents a policy that prohibits people from leaving their home regardless of their testing status. For some countries, the policy can also include the case when people have to stay at home, but are allowed to leave for work- or health-related purposes. For the latter case, when the policy is moderate, this is coded as *home isolation = 0*.*5*.
- *work from home* : Represents a policy that requires people to work remotely. This policy may also include encouraging workers to take holiday/paid time off.
- *business closure* : Represents a policy that closes all offices, non-essential businesses, and non-essential commercial activities in that area.
- *pos cases quarantine* : A policy that mandates that people who have tested positive for COVID-19, or subject to quarantine measures, have to confine themselves at home. The policy can also include encouraging people who have fevers or respiratory symptoms to stay at home, regardless of whether they tested positive or not.
- *welfare service closure*: A policy that mandates closure of welfare services such as day care centers for children.
- *emergency declaration*: Represents a decision made at the city / municipality, county, state / provincial, or federal level to declare a state of emergency. This allows the affected area to marshal emergency funds and resources as well as activate emergency legislation.

##### iv. Social Distance

- *social distance*: Represents a policy that encourages people to maintain a safety distance (often between one to two meters) from others. This policy differs by country, but includes other policies that close cultural institutions (e.g., museums or libraries), or encourage establishments to reduce density, such as limiting restaurant hours.
- *no gathering* : Represents a policy that prohibits any type of public or private gathering. (whether cultural, sporting, recreational, or religious). Depending on the country, the policy can prohibit a gathering above a certain size, in which case the number of people is specified by the *no gathering size* variable.
- *event cancel* : Represents a policy that cancels a specific pre-scheduled large event (e.g., parade, sporting event, etc). This is different from prohibiting all events over a certain size.
- *religious closure*: Represents a policy that prohibits gatherings at a place of worship, specifically targeting locations that are epicenters of COVID-19 outbreak. See the section on Korean policy for more information on this policy variable.
- *no demonstration*: Represents a policy that prohibits protest-specific gatherings. See the section on Korean policy for more information on this policy variable.

#### A.3 Mobility data

Mobility data comes from three of the biggest internet companies - Google, Facebook and Baidu. These companies have millions of users accessing their social media, e-commerce and other digital platforms every day. These data are utilized to construct aggregated, anonymized user location and movement metrics for various geographic regions and countries. Descriptions follow, and Table 3 contains a summary of the data used for each country.

##### A.3.1 Google

Google mobility data summarizes time spent by their users each day after Feb 6, 2020 in various types of places, such as residential, workplaces and grocery stores.^11^ Specifically, it provides the percentage change in number of visits and length of stay in each type of place, compared to a baseline value. The baseline is the value on the corresponding day of the week during the 5-week period between Jan 3, 2020 and Feb 6, 2020. The metrics are available starting Feb 15, 2020 at the country (Administrative Level 0) and state level (Administrative Level 1) for over 135 countries. We also access county-level metrics (Administrative Level 2) for the US. Types of places include the following:

i. *Grocery & pharmacy* : Places like grocery markets, food warehouses, farmers markets, specialty food shops, drug stores, and pharmacies.
ii. *Parks*: Places like local parks, national parks, public beaches, marinas, dog parks, plazas, and public gardens.
iii. *Transit stations*: Places like public transport hubs such as subway, bus, and train stations.
iv. *Retail & recreation*: Places like restaurants, cafes, shopping centers, theme parks, museums, libraries, and movie theaters.
v. *Residential* : Places of residence.
vi. *Workplaces*: Places of work.

##### A.3.2 Facebook

Facebook summarizes and anonymizes its user data into useful metrics that can be used to evaluate the movement of people.^12,13^ Our analysis uses data beginning March 5, Feb 23 and Feb 24, 2020 for France, Italy and South Korea respectively. Specifically, Facebook aggregates the number of trips between tiles of up to a resolution of 360 square meters. We aggregate these data to the level of administrative regions, constructing metrics for number of trips *between* as well as *within* these regions. We use the following variables from the data provided by Facebook:

i. *Date* - The day of the movement.
ii. *Starting Location* - The region or tile where the movement of the group started.
iii. *Ending Location* - The region or tile where the movement of the group ended.
iv. *Baseline Movement* - The total number of people who moved from Starting Location to Ending Location on average during the weeks before the disaster began.
v. *Crisis Movement* - The total number of people who moved from Starting Location to Ending Location during the time period specified

##### A.3.3 Baidu

Baidu provides aggregated user location data and mobility metrics via its Smart Eye Platform.^14^ These data were scraped and publicly shared by the China Data Lab. The metrics represent movement in and out of major regions across China each day in terms of an aggregated mobility index (China-Data-Lab, 2020). Index values are available beginning Jan 1, 2020. Baidu does not disclose specific information regarding the construction of the index.

##### A.3.4 SafeGraph

SafeGraph data were generated by tracking anonymous mobile devices across US.^15^ The mobility metrics are available starting January 1, 2020 for census block group. SafeGraph infers home location based on night time location of the device and uses that to impute average distance travelled per day by the devices in each census block. We aggregate this data to the state level (Administrative Level 1) for our analysis.

### B Methods Summary

#### B.1 Behavior Model

The behavior model describes how human mobility changes as a result of NPIs 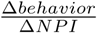 in equation (1)). The model is a commonly used reduced-form approach in econometrics. Details on the model and model estimation are presented below.

##### Model details

1. The model used for each policy is *m*_*t*_ = *f* (*policy*_*t*_, *X*_*t*_) + *ϵ*_*t*_, where *m*_*t*_ is a measure of mobility behavior at time *t, X*_*t*_ represents control variables, and *E*_*t*_ is the error.
2. The model is fit for each country at the sub-national level where granular policy and mobility data are available. For the rest of the world, use a panel regression model where the unit of observation is at the country by day level.
3. The *policy* variable is a vector with NPIs specific to each country, for each location and day. NPIs are continuous variables between 0 and 1 (inclusive) that indicate the intensity of the policy where 0 is no enforcement and 1 is fully enacted. In some instances, it may be desirable to gather multiple policies in a single variable (for example, business closure and restaurant closure) by taking the average, thus the maximum value of 1 would indicate that all policies are fully enacted.
4. The control variable *X* includes one-hot encodings of sub-national (or national) units and day-of-week variables. The former account for time-invariant factors (for example, socio-economic status, culture, public transportation availability) that impact mobility *m*, while the later control for weekly patterns in mobility (for example, less workplace related mobility on Sunday) that are common across location unit.

##### Steps for model estimation

1. Estimate the average effect on mobility in all subsequent periods, 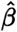, of each policy included in each model using the model described above, and ordinary least squares.
2. Compute the combined effect of policies on human mobility by taking the sum across all 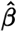.

#### B.2 Infection Model

Similar to the behavior model, the infection model is also a reduced-form approach, used to describe the relationship between infections and mobility behavior (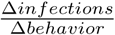 in equation (1)). Model details, as well as steps for model estimation, forecasting and cross-validation are outlined below. Also included are steps for data selection.

#### Model details

1. The model used is 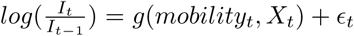, where 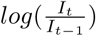 is the first-difference of log confirmed infections at time *t, X*_*t*_ represents control variables, and *ϵ*_*t*_ is the error.
2. The model is fit for each country at the sub-national level where granular infections and mobility data are available. For the global model, use a regression model where the unit of observation is at the country by day level.
3. The *mobility* variable is a vector with mobility rates specific to each country, for each location and day. Includes mobility measures averaged over lags 1-7, 8-14 and 15-21, respectively. We use Google mobility data in its original form (percentage points), and take logs for the Facebook and Baidu mobility data.
4. The control variable *X* includes one-hot encodings of sub-national (or national) units and day-of-week variables.

##### Steps for model estimation

The following steps are used to generate estimates of the average effect of each mobility variable on the growth rate of infections (see Figure 6). These are then used to estimate how a novel policy affecting mobility would alter future infections (Table 2).

1. Estimate the average effect of each mobility variable on the growth rate of infections, 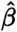, using the model described above, and ordinary least squares.
2. To estimate the potential effect of a mobility-reducing policy, use 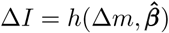, where Δ*I* is the change in number of infections, Δ*m* is the anticipated change in mobility due to NPIs, and 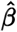 are estimated coefficients of the mobility variables.
3. Specifically, for Facebook and Baidu data, where mobility variables are in log form: at forecast day 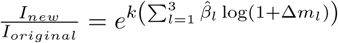, where Δ*m* is the fractional change in the *l*th mobility variable (number of trips for all lags involved) (e.g., if the number of trips for all lags in the *l*th variable is reduced by 10%, Δ*m*_*l*_ = −.1). For Google data: at forecast day 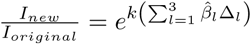, where Δ*m* is the change in residential time over baseline for the *l*th mobility variable (e.g., Δ*m*_*l*_ = .05 means a 5% increase, say from 20% to 25% residential time over baseline, for all lags in the *l*th variable).

#### Steps for forecasting and cross-validation

1. For a 20-day period (training data), fit a regression model as specified above, using ordinary least squares.
2. For a 10-day period (test data), multiply the coefficient estimates obtained from fitting the regression model on the training data with the observed predictor variables in the test data to obtain prediction of the infection rate.
3. For each test day, compute the percentage error compared to the ground-truth infection rate.
4. Perform cross-validation (i.e., robustness to train and test sample selection), for all 20-day training periods and 10-day forecast periods, limited by data availability.
5. Group percentage errors by day of forecast (from 1 to 10).
6. Repeat the above, using a baseline model which excludes mobility variables, i.e., 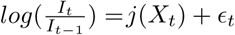.

In other words, the baseline model simply uses past infections to predict future infections. Smaller errors using the model including mobility variables would indicate that information on mobility improves forecasts. Examples of the results are in Figure 3, and forecasting errors are in 4.

#### Steps for data selection

The time period that we consider (see Table 3) is the “first wave” of infections, and to demonstrate the utility of the mobility model, we focus on the period in which mobility starts falling as a result of lockdown measures imposed during this first wave, until right before mobility starts increasing again. This model can be refit to the local context of interest, using data that is representative of current conditions. For countries in which lockdown policy data are available, we include administrative regions after the lockdown policy has been implemented. For countries without policy data available on a granular enough level (US in this case), we use a start date of March 17 (the results are robust to different start dates), or when Google residential mobility is at least 5 percent above baseline (world level). We select an end date that roughly corresponds to just before mobility picks back up. The reason for this choice is that in the phase in which mobility starts to increase, we might expect there to be other measures put in place, or other changes in behavior, such as contact tracing, mask wearing, and so forth, which justified the lifting of lockdown measures and subsequent increase in mobility. The relationship between mobility and cases might therefore be different than during the lockdown stage, suggesting that the model needs to be refit if we would like for it to be used during this period.

Now, we train each model using 20 days of training data, and forecast for up to 10 days into the future. To be included in the data used to train each model, we impose the following conditions at the level of the *administrative region*. For the administrative region to be included, for *all* 20 training days *t*,

1. *I*_*t*_ *≥* 10
2. *I*_*t*−1_ *>* 0

and for the world-level analysis only:

1. mobility_*t*_ *≥* 5 percent, i.e., current day mobility is at least 5 percent above baseline.
2. 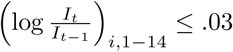 percent, i.e., the 14 days rolling average of the growth rate of cumulative active cases flattens.

These conditions also imply that *I*_*t*_, *I*_*t*−1_ and the mobility variables have to be non-missing for all training days. These conditions have implications for predictions as well: if an administrative region is not included in the training data, predictions will not be generated for that region, because the region fixed effect would not be estimated for that region.

### C. Additional Figures

## Footnotes

1 Global Policy Lab, UC Berkeley, http://www.globalpolicy.science/covid19, website accessed on October 20, 2020.

2 The Organisation for Economic Co-operation and Development, https://www.oecd.org/coronavirus/en/#country-tracker, website accessed on April 12, 2020.

3 Kaggle, COVID-19 lockdown dates by country, https://www.kaggle.com/jcyzag/covid19-lockdown-dates-by-country, website accessed on April 12, 2020.

4 Google, COVID-19 Community Mobility Reports, https://www.google.com/covid19/mobility/, website accessed on March 20, 2020.

5 Facebook Disaster Maps: Aggregate Insights for Crisis Response and Recovery, https://research.fb.com/publications/facebook-disaster-maps-aggregate-insights-for-crisis-response-recovery/, website accessed on March 20, 2020.

6 Baidu, Spatio-temporal Big Data Service, https://huiyan.baidu.com, website accessed on March 20, 2020.

7 SafeGraph, Social Distancing Metrics, https://docs.safegraph.com/docs/social-distancing-metrics, website accessed on March 20, 2020.

8 COVID-19 Data Repository by the Center for Systems Science and Engineering (CSSE) at Johns Hopkins University, https://github.com/CSSEGISandData/COVID-19, website accessed on March 20, 2020.

9 https://github.com/CSSEGISandData/COVID-19

10 http://www.globalpolicy.science/covid19

11 https://www.google.com/covid19/mobility/

12 https://research.fb.com/publications/facebook-disaster-maps-aggregate-insights-for-crisis-response-recovery/

13 https://about.fb.com/news/2017/06/using-data-to-help-communities-recover-and-rebuild/

14 https://huiyan.baidu.com

15 https://docs.safegraph.com/docs/social-distancing-metrics

